# Mouth-rinses and SARS-CoV-2 viral load in saliva: A living systematic review

**DOI:** 10.1101/2021.03.23.21254214

**Authors:** Akram Hernández-Vásquez, Antonio Barrenechea-Pulache, Daniel Comandé, Diego Azañedo

## Abstract

**Objective:** To conduct a living systematic review of the clinical evidence regarding the effect of different mouth-rinses on the viral load of SARS-CoV-2 in the saliva of infected patients. The viral load in aerosols, the duration of the reduction in viral load, viral clearance, SARS-CoV-2 cellular infectivity, and salivary cytokine profiles were also evaluated.

**Materials and methods:** This study was reported using the PRISMA guidelines. An electronic search was conducted in seven databases and in preprint repositories. We included human clinical trials that evaluated the effect of mouth-rinses with antiseptic substances on the viral load of SARS-CoV-2 in the saliva of children or adults that tested positive for SARS-CoV-2 using reverse transcriptase polymerase chain reaction (RT-PCR). Risk of bias was assessed using the ROBINS-I tool. PROSPERO registration number CRD42021240561.

**Results:** Four studies matching eligibility criteria were selected for evaluation (n=32 participants). Study participants underwent oral rinses with hydrogen peroxide (H2O2) at 1 %, povidone–iodine (PI) at 0.5% or 1%, chlorhexidine gluconate (CHX) at 0.2% or 0.12% or cetylpyridinium chloride (CPC) at 0.075%. Only one study included a control group with sterile water. Three of the studies identified a significant reduction in viral load up to 3, 4, and 6 hours after the use of mouthwashes with PI, CHX, and CPC or PI vs. sterile water, respectively, while one study did not identify a significant reduction in viral load after the use of H2O2 rinses.

**Conclusions:** According to the present systematic review, the effect of the use of mouth-rinses on SARS-CoV-2 viral load in the saliva of COVID-19 patients remains uncertain. This is mainly due to the limited number of patients included and a high risk of bias present in the studies analyzed. Evidence from well-designed randomized clinical trials is required for further and more objective evaluation of this effect.

## INTRODUCTION

The SARS-CoV-2 pandemic has had a great impact on the health of the global population due to its increasing spread and lack of treatment to prevent infection or reduce disease severity. Since the first case report in November 2019 (Kpozehouen et al. 2020) and up to March 2021, more than 121 million infections and over 2.6 million deaths have been registered and cases continue to increase (Dong, Du, Gardner 2020). The global scientific community has therefore started a series of studies aimed at identifying public health measures and technologies to mitigate the spread of this disease.

So far, the only effective measures endorsed by the World Health Organization (WHO) to control and prevent the transmission of this disease at the community level are continuous hand washing, physical distancing, and the correct use of face masks (World Health Organization 2021). Likewise, in relation to hospital care, the WHO has recommended the use of personal protective equipment (glasses / face shields, gloves, gowns, and N95 respirators or their equivalents), for health personnel who care for patients with COVID-19 and perform activities with a high risk of contagion, such as aerosol-generating procedures performed by dentists. These procedures can cause viral shedding through aerosols and splashes generated during dental care, potentially infecting health care personnel or patients seen between appointments.

Since there is no known treatment for COVID-19, the administration of a series of drugs currently being tested in clinical trials or approved for other uses has been proposed for off-label use during the pandemic, as preventive or recuperative therapies for COVID-19 based on studies describing some level of *in vitro* activity against SARS-CoV-2. Nonetheless, clinical studies published later have shown a lack of efficacy of these drugs to prevent or treat the disease, and in some cases, they were associated with higher mortality and morbidity (Ghazy et al. 2020). In the field of dentistry, various investigators have proposed that the pre-procedural use of mouth-rinses that include antiseptic substances could generate a reduction of SARS-CoV-2 viral load in the saliva of infected patients (O’Donnell et al. 2020; Peng et al. 2020). These proposals are mainly based on evidence from *in vitro* studies (Meister et al. 2020), and more recently, human clinical trials (Seneviratne et al. 2020).

In response, international organizations such as the *Centers for Disease Control and Prevention* and the *American Dental Association* (American Dental Association 2020; Center for Disease Control and Prevention 2020) have incorporated recommendations about the use of mouth-rinses before dental procedures in their guidelines for dental health care during the COVID-19 pandemic. These recommendations are aimed at reducing the load of oral microorganisms in aerosols generated during treatment, emphasizing the lack of clinical evidence about their efficacy in reducing the transmission of SARS-CoV-2. This has generated controversy among the population, mouth-rinse manufacturing companies, and health care professionals and researchers with regard to the applicability of this measure in the prevention against SARS-CoV-2 infection (Reis et al. 2021).

Due to the current interest in the utility of this oral hygiene product in the prevention of SARS-CoV-2 infection, the objective of this study was to make a living systematic review of the clinical evidence regarding the effect of different mouth-rinses on the viral load of SARS-CoV-2 in the saliva of infected patients. Additionally, we aimed to evaluate the effect of mouthwashes on the quantification of the virus in the aerosols generated during dental care of infected patients, and the duration of the reduction of viral load in saliva and aerosols. We will also perform periodic updates of the study search, since according to the ClinicalTrials.gov registry, clinical trials on the subject are ongoing (Alzahrani 2021; Jacox 2021; Villar 2021), and will eventually provide new clinical evidence to complement the currently published information and the conclusions of this systematic review.

## MATERIALS AND METHODS

The protocol of this living systematic review was prospectively registered in accordance with the Preferred Reporting Items for Systematic Reviews and Meta-Analyses Protocols (PRISMA-P) statement (Moher et al. 2015) on PROSPERO with reference number CRD42021240561. We will update the search on a monthly basis according to the availability of new evidence as a living systematic review (Elliott et al. 2017) and we plan to maintain the review in a living mode for at least 12 months from the publication of the protocol. This article currently reports the basal findings from relevant articles identified up to February 26th, 2021. The reporting of the results complies with the ‘Preferred Reporting Items for Systematic reviews and Meta-Analyses’ (PRISMA) guidelines (Moher 2009) (Appendix Checklist 1).

## Inclusion Criteria

We included human clinical trials that evaluated the effect of mouth-rinses with antiseptic substances on the viral load of SARS-CoV-2 in the saliva of patients, children, or adults with COVID-19, diagnosed using reverse transcriptase polymerase chain reaction (RT-PCR). Likewise, we included studies that evaluated the outcome of viral load on aerosols generated during dental procedures, the duration of the reduction in viral load, viral clearance, SARS-CoV-2 cellular infectivity in saliva, and salivary cytokine profiles. We did not restrict our criteria to any dosage, duration, or timing of mouth-rinses. The comparison of interest was distilled water, sterile water, tap water, saline solution, or no treatment. We included articles published in both peer-reviewed journals and preprints and only publications written in English or Spanish. We excluded comments, conference abstracts, interviews, and studies developed in animal models or *in vitro* conditions. The preprints included will be reassessed at the time of peer-reviewed publication, and the most current version will be included.

## Search strategy

A systematic electronic search of articles published until February 26th, 2021 was conducted using PubMed, CINAHL, The Cochrane Library, Embase, Scopus, Dentistry & Oral Sciences Source, and LILACS databases. Preprint repositories including medRxiv and bioRxiv were also searched.

A librarian (DC) developed the search strategies which were later validated by the other authors (AHV, ABP and DA). We initially designed a search strategy in PubMed which was adapted to the other databases containing the following terms with synonyms and other medical descriptors: «coronavirus», «SARS-CoV-2», and «Mouth-rinse». The details of the search terms used for the search are shown in Appendix Table 1. The same search strategies will be run on every update. We also manually screened the references of the original studies and reviews included to identify additional eligible studies. No date, language, study design, publication status, or language restriction was applied to the searches in the databases.

**Table 1.** Characteristics of the studies included.

| Author/Year | Type of publication | Country | Setting | Sample (n) | COVID-19 Diagnosis | Mouth-rinse type | Experiment details | Primary Outcome evaluated | Follow-up (time) |
| --- | --- | --- | --- | --- | --- | --- | --- | --- | --- |
| Gottsauner (2020) | Short communication | Germany | Hospital | 10 | Only patients with a positive RT-PCR test for SARS-CoV-2 within the last 72 h were included in this study | 20 mL 1% H2O2 | Patients were asked to gargle their mouth and throat with 20 mL 0.9% NaCl for 30 s in order to acquire a baseline oropharyngeal specimen for the SARS-CoV-2 real-time RT-PCR test. Immediately afterwards, patients had to perform a mouth rinse with 20 mL 1% H2O2 by gargling their mouth and throat for 30 s. Thirty minutes after this mouth rinse, another oropharyngeal specimen for the SARS-CoV-2 RT-PCR test was acquired by letting the patients gargle their mouth and throat with 20 mL 0.9% NaCl for 30 s. The respective quantities of copies/mL of SARS-CoV-2 RNA were analyzed by RT-PCR. | Intraoral viral load in SARS-CoV-2-positive patients | 30 minutes |
| Martínez Lamas (2020) | Short communication | Spain | Hospital | 4 | RT-PCR assay | 15 ml of 1% PI | Patients were asked to cough out saliva from their throat into a sterile container, and 2 mL of viral transport medium was added at baseline after which each participant rinsed their mouth with 15 ml of 1% PI for 1 min. Afterwards saliva samples were collected at 5 min, 1h, 2h and 3 hours. | Salivary viral load of SARS-CoV-2 in 4 patients with COVID-19 | 3 hours |
| Seneviratne (2020) | Original article | Singapore | Hospital | 16 | Positive nasal swabs for SARS-CoV-2 based on RT-PCR assay | 1. PI 5 ml (0.5% w/v)<br>2. CHX 15 ml of undiluted (0.2% w/v)<br>3. CPC 20 ml of 0.075% | Three mL of saliva was collected by passive drool technique from all the enrolled COVID-19 patients at baseline and after rinsing their mouth with the corresponding mouthwash (PI, CHX, CPC or water). Three additional samples were obtained at 5 min, 3 hours and 6 hours. | Salivary SARS-CoV-2 viral load | 6 hours |
| Yoon (2020) | Original article | Korea | Hospital | 2 | Real-time RT-PCR | CHX (0.12%, 15 mL) for 30 seconds | During hospital days 3 and 6, patients were asked to collect saliva samples while being supervised by<br>an investigator in order to obtain a baseline for the SARS-CoV-2 using real-time RT-PCR test. Immediately afterwards, patients had to perform a mouth rinse with CHX solution mouth-rinse (0.12%, 15 mL) for 30 seconds. Serial saliva samples were collected at 1, 2 and 4 hours after the mouth-rinse for the SARS-CoV-2 real-time RT-PCR test. The respective quantities of copies/mL of SARS-CoV-2 RNA were analyzed by real-time RT-PCR. | Viral dynamics in various body<br>fluid specimens, such as nasopharyngeal swab, oropharyngeal swab, saliva, sputum, and urine specimens, of two patients with COVID-19 from hospital day 1 to 9 | 4 hours |

RT-PCR: reverse-transcription polymerase chain reaction.
H2O2: hydrogen peroxide; PI: povidone–iodine; CHX: chlorhexidine gluconate; CPC: cetylpyridinium chloride; NaCl: Sodium Chloride.

In this living systematic review, three independent authors (DC, ABP, and AHV) will receive an updated literature search file every month. They will continuously include relevant newly published or unpublished studies as per the above inclusion criteria.

## Study selection

Initially, the results of the electronic search were imported to the reference management software EndNote X9. We eliminated all duplicate registries following the methodology described by Bramer *et al*. (Bramer et al. 2016). An independent two-stage screening process was undertaken to identify studies meeting the eligibility criteria by two authors (ABP and AHV) using the web application “Rayyan” (Ouzzani et al. 2016). First, we evaluated the registries by title and abstract, and those appearing to meet the inclusion criteria were selected and the remaining were discarded. Afterwards, the full texts of the selected papers were evaluated under the same inclusion criteria. Any disagreement was discussed among the reviewers and in case of not being able to make a decision a third reviser took part in the discussion (DA).

## Extraction and synthesis of results

The report of the reduction of viral load of SARS-CoV-2 in saliva or aerosols generated by dental procedures, and the duration of said reduction, was planned, using the different units of measurement registered before and after the use of mouth-rinses according to the report of each study. We also extracted the following information: study design, settings, participant characteristics, study eligibility criteria, intervention, control, and the risk of bias assessment for each study. Data was independently extracted by two authors (ABP and AHV) using standardized forms. If these data were not reported, we contacted authors to request them.

The general information about the publications and specific data of each study included was compiled in the summary tables. For any outcome in which data were sufficient to calculate an effect estimate, we planned to conduct a meta-analysis.

## Risk of Bias Assessment

Two authors (AHV and ABP) independently assessed the risk of bias using the ROBINS-I (Risk Of Bias In Non-Randomized Studies of Interventions) tool (Sterne et al. 2016) and disagreements were resolved through discussion with a third author (DA).

## Ethical considerations

We did not solicit the approval of the study by an Institutional Review Board because this is a revision of bibliographic databases.

## RESULTS

The search strategy among the different databases identified 619 articles. After removing duplicates 381 articles were included, and a total of 373 studies were excluded in phase one after review by title and abstract. The remaining eight articles were evaluated in full text, and four were excluded after this evaluation; the reasons for exclusion were: not measuring viral load in saliva or aerosols after the intervention with mouth-rinses (three studies), and only considering qualitative measurements to establish the diagnosis of COVID-19 (one study) (Figure 1). After this process, four studies (Appendix References 1) were included (Gottsauner et al. 2020; Martínez Lamas et al. 2020; Seneviratne et al. 2020; Yoon et al. 2020).

**Figure 1.**
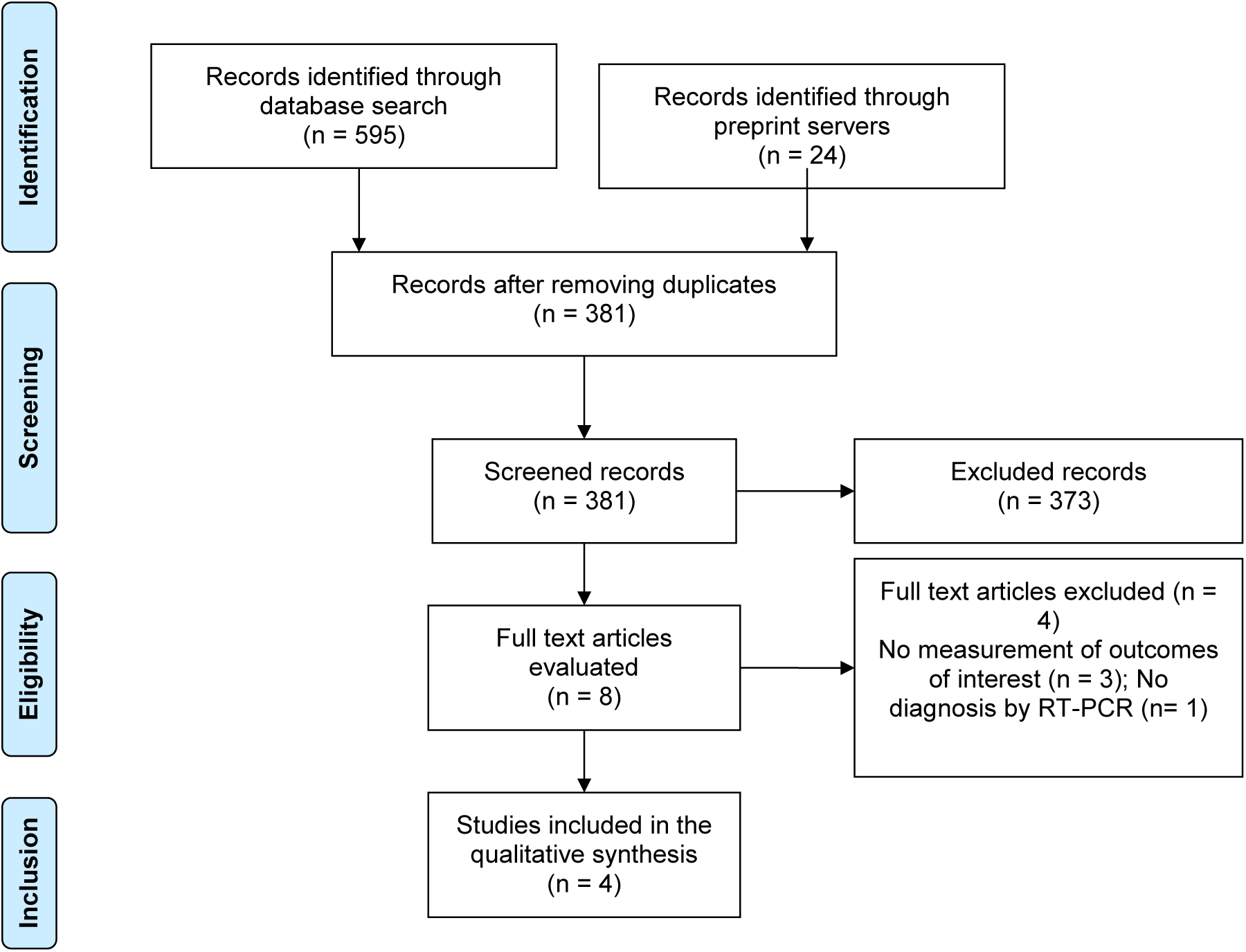
Flow chart of study selection according to the PRISMA statement.

**Figure 2.**
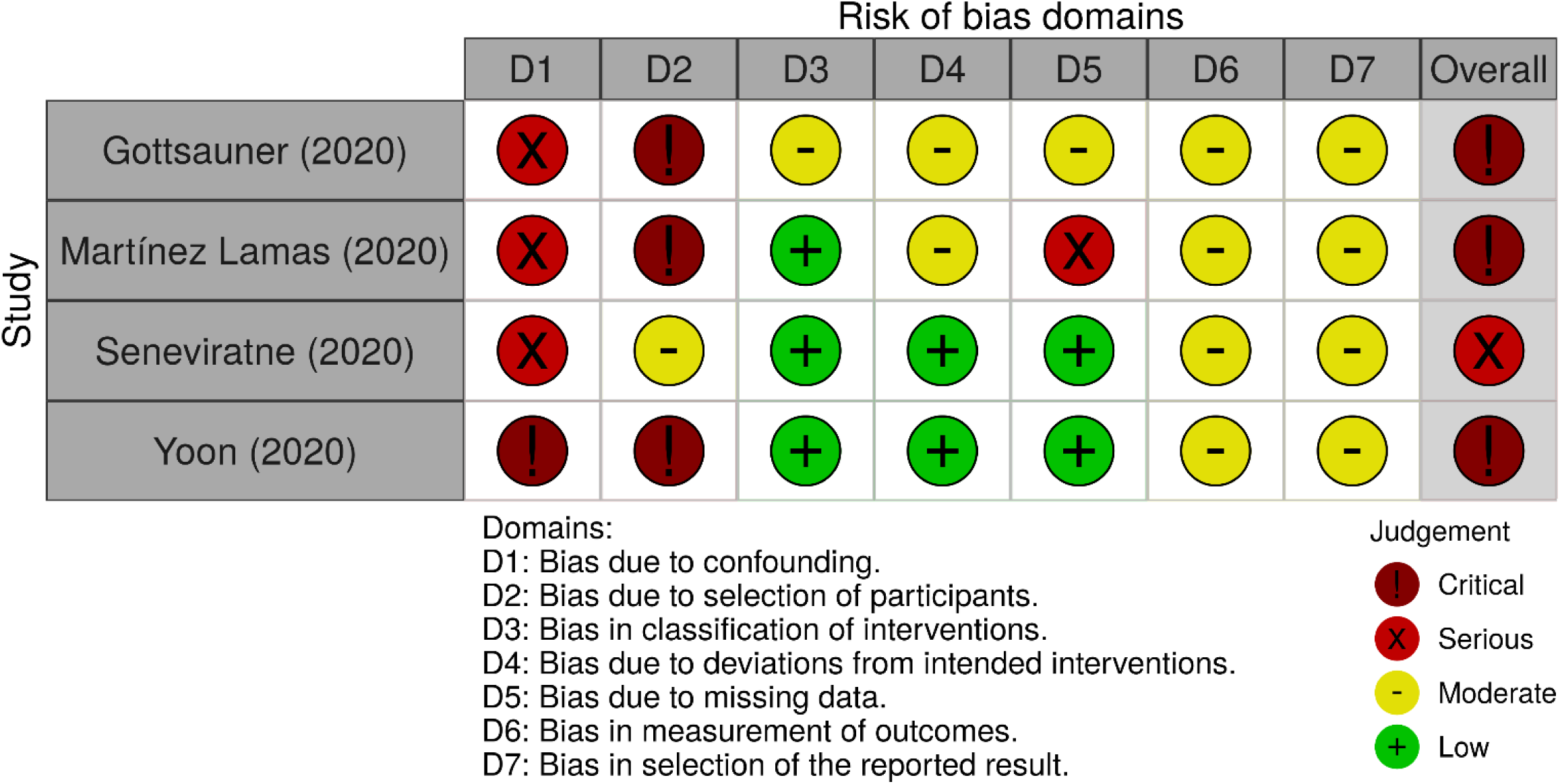
Risk of bias using ROBINS-I.

Half of the studies included were original articles (Seneviratne et al. 2020; Yoon et al. 2020), while the rest were short communications (Gottsauner et al. 2020; Martínez Lamas et al. 2020). All were reported in English (n=4). Among the four studies a total of 32 participants were included. The patients subjected to an intervention received mouth-rinses with either hydrogen peroxide (H2O2) 1%, povidone–iodine (PI) (0.5% or 1%), chlorhexidine gluconate (CHX) (0.2% or 0.12%) or cetylpyridinium chloride (CPC) (0.075%). Only one study (Seneviratne et al. 2020) included a control group that used sterile water for comparisons. This study included the largest number of participants (n=16) and was also the only study to mention having utilized commercially available mouth-rinses with PI, CHX and CPC (Table 1).

The main objective of these studies was to evaluate the effect of the use of different mouth-rinses on the viral load of SARS-CoV-2 in the oral cavity or saliva. Measurements of this outcome were conducted before and after the intervention at variable time intervals. The minimum interval of time post-intervention for the first measurement was 5 minutes, and the maximum was 6 hours. The highest number of measurements per participant was 5 (basal, after 5 minutes, 1 hour, 2 hours, and 3 hours) and the lowest was 2 (basal and after 30 minutes). Most studies were carried out in a single day and only one conducted serial measurements at the hospital on days 3 and 6 (Yoon et al. 2020) (Table 1).

The viral load obtained during follow-up was reported as copies/mL and only one study reported it as cycle threshold value. Measurements of viral load were highly variable and at times conflicting among the studies (Table 2). Yoon (2020) observed in his intervention on 2 patients during hospital day three, that after use of mouth-rinses based on CHX the viral load became undetectable during 2 and 4 hours of follow-up, becoming detectable again after 6 hours. Nonetheless, on hospital day 6 the viral load of SARS-CoV-2 was detectable throughout all measurements of follow-up (Yoon et al. 2020). Meanwhile, Seneviratne (2020) identified statistically significant differences in viral load after the use of mouth-rinses in comparisons between CPC vs. water at 5 minutes and 6 hours of follow-up, and PI vs. water at 6 hours of follow-up (Seneviratne et al. 2020). Likewise, Martínez Lamas (2020) observed a progressive reduction of the mean viral load obtained from the four participants from baseline until 3 hours of follow-up (Martínez Lamas et al. 2020). These three authors agree that the use of mouth-rinses could reduce the viral load of SARS-CoV-2 in the saliva samples of patients, reaffirming the need for more thorough studies to confirm these preliminary findings (Martínez Lamas et al. 2020; Seneviratne et al. 2020; Yoon et al. 2020). In contrast, Gottsauner (2020) concluded that mouth-rinses based on H2O2 1% did not reduce the intraoral viral load of those infected with the new coronavirus and was not able to establish a significant reduction in the viral load in saliva after 30 minutes of follow-up in 10 patients (Gottsauner et al. 2020).

**Table 2.** Summary of results of the studies.

| Author/Year | Sample (n) | Intervention | Comparator | Results for primary outcome (viral load) |
| --- | --- | --- | --- | --- |
| Gottsauner (2020) | 10 | 20 mL 1% CHX | None | Baseline: Median $1.8 \times 10^3$ (IQR $3.1 \times 10^2$ and $4.7 \times 10^4$) copies/mL of SARS-CoV-2 RNA<br>30 min: Median $1.5 \times 10^3$ (IQR $8.3 \times 10^2$ and $3.4 \times 10^4$) copies/mL of SARS-CoV-2 RNA |
| Martínez Lamas (2020) | 4 | 15 ml of 1% PI | None | Baseline: mean 3.75 log<sub>10</sub> copies/mL<br>5min: mean 3.9 log<sub>10</sub> copies/mL<br>1h: mean 2.72 log<sub>10</sub> copies/mL<br>2h: mean 2.27 log<sub>10</sub> copies/mL<br>3h: mean 2.15 log<sub>10</sub> copies/mL |
| Seneviratne (2020) | PI group (n=4)<br>CHX group (n=6)<br>CPC group (n=4)<br>water as control group (n=2) | 1. PI 5 ml (0.5% w/v)<br>2. CHX 15 ml of undiluted (0.2% w/v)<br>3. CPC 20 ml of 0.075% | Sterile water | Baseline: Cycle Threshold value PI group 22<br>CHX group 30<br>CPC group 33<br>water group 26<br>5 min: Cycle Threshold value PI group 24<br>CHX group 28<br>CPC group 34<br>water group 25 --- CPC vs. water $p < 0.05$<br>3 h: Cycle Threshold value PI group 24<br>CHX group 30<br>CPC group 31<br>water group 24<br>6 h: Cycle Threshold value PI group 22<br>CHX group 28<br>CPC group 32<br>water group 23 --- CPC vs. water $p < 0.05$, PI vs water $p < 0.01$ |
| Yoon (2020) | 2 | CHX solution mouthwash (0.12%, 15 mL) for 30 seconds | None | Hospital Day 3:<br>Baseline: mean 5.865 log<sub>10</sub> copies/mL<br>1 h: non detectable<br>2 h: non detectable<br>4 h: mean 3.39 log<sub>10</sub> copies/mL<br>Hospital Day 6<br>Baseline: mean 4.68 log<sub>10</sub> copies/mL<br>1 h: mean 4.315 log<sub>10</sub> copies/mL<br>2 h: mean 3.39 log<sub>10</sub> copies/mL<br>4 h: mean 4.495 log<sub>10</sub> copies/mL |

IQR: interquartile range; PI: povidone-iodine; CHX: chlorhexidine gluconate; CPC: cetylpyridinium chloride.

## Assessment of risk of bias

After using the ROBINS-I tool we concluded that the four studies analyzed have a critical (Gottsauner et al. 2020; Martínez Lamas et al. 2020; Yoon et al. 2020) or serious risk (Seneviratne et al. 2020) of bias mainly due to not controlling for confounding factors and the selection of patients. Regarding the first domain, the majority of studies did not take into account the beginning of COVID-19 symptoms, did not control the ingestion of food or water by patients prior to the start of the intervention, and in one study participants had been taking antiviral medication during the duration of the study (Yoon et al. 2020). For the second domain, these studies had very small samples, with the largest including only 16 patients (Seneviratne et al. 2020). Moreover, the characteristics of the participants were very heterogeneous between the studies and, furthermore, the studies did not clearly establish inclusion or exclusion criteria.

## DISCUSSION

The main objective of this living systematic review was to evaluate the effect of different kinds of mouth-rinses on the viral load of SARS-CoV-2 in the saliva of patients with COVID-19. Four studies were included; two published in original article format and two as short communications. They included patients with positive SARS-CoV-2 infection confirmed by RT-PCR (32 participants in total), who underwent oral rinses with H2O2 at 1 %, PI at 0.5% or 1%, CHX at 0.2% or 0.12% or CPC at 0.075%. Only one of the studies included a control group in which sterile water was used. All studies included a baseline assessment of viral load in saliva or oropharyngeal secretions and between two and five follow-up periods with a minimum of five minutes and a maximum of six hours after the intervention. Measurement of viral load was reported in copies / mL and one of the studies used the cycle threshold value to determine a detectable viral load. Three of the studies identified a significant reduction in viral load up to 3, 4, and 6 hours after the use of mouth-rinses with PI, CHX, and CPC or PI vs. sterile water, respectively, while one study did not identify a significant reduction in viral load after the use of H2O2 rinses. Likewise, the four studies evaluated were at high risk of bias.

The results of three studies reported that the use of mouth-rinses with PI, CHX, or CPC could reduce the viral load of SARS-CoV-2 in the saliva of confirmed COVID-19 patients. However, it should be taken into account that the evidence to date corresponds to studies with a small sample size (between 2 and 16 patients), with a risk of serious or critical bias, mainly due to the impossibility of controlling the potential effect of confounding variables. For example, the studies did not take into account variables such as the onset of COVID-19 symptoms or the consumption of food prior to the intervention, which could have an impact on the viral load in saliva. Therefore, with the results of these studies, it is not possible to conclude that the reduction in viral load is due solely to the effect of mouth-rinses. In addition, despite three of the four studies included showing a positive effect on reducing the viral load of SARS-CoV-2, we could not say that these results are consistent, due to the marked heterogeneity among the studies. These differences were mainly identified in the measurement units used to express viral load, the number and periods of follow-up, the concentrations of antiseptic substances, and the procedures for the use of mouth-rinses, among other aspects.

On the other hand, most of the studies analyzed suggest that different types of mouth-rinses reduce the viral load of SARS-CoV-2 in the saliva of infected patients up to six hours after their application. This could translate into a reduction in the viral content found in aerosols generated during dental procedures, resulting in a reduction in the spread of the disease in health personnel and patients without infection. However, post-intervention measurements in the studies included were not performed after the performance of a dental procedure. This is important because it is hypothesized that SARS-CoV-2 would be present in the salivary glands of infected patients (Xu, J. et al. 2020; Xu, R. et al. 2020), being expelled into the oral cavity through saliva (Sakanashi et al. 2021; Wyllie et al. 2020; Zhu et al. 2020). So, given that salivary secretion is stimulated during oral instrumentation for dental procedures, it would, therefore, be reasonable to conclude that the duration in the reduction of viral load would probably be of shorter in clinical scenarios. Evidence from studies of a higher methodological level, conducted in real or simulated clinical settings are of vital importance to better understand the effect of mouth-rinses on SARS-CoV-2 viral load in the saliva of patients with COVID-19.

One of the limitations of the study is that due to the low quality of the studies, small sample size, as well as the heterogeneity reported in terms of follow-up periods, and reporting of viral load measurement, the calculation of a meta-analysis would have very limited utility. However, an extensive bibliographic search has been carried out in seven databases, and the search has also been extended to preprint repositories including medRxiv, and bioRxiv, in order to cover the greatest amount of evidence available on the subject. Likewise, the present study has a living systematic review design in order to perform future updates of this study according to the results of completed or ongoing clinical trials registered in the ClinicalTrials.gov portal. This will enable the adoption of a more conclusive position for or against the use of mouth-rinses in reducing the viral load of SARS-CoV-2 in the saliva of patients with COVID-19.

In conclusion, according to the present systematic review, the effect of the use of mouth-rinses on the viral load of SARS-CoV-2 continues to be uncertain. The emission of recommendations by governmental organizations around the world about the use of these mouth-rinses as a preventive measure towards infection by SARS-CoV-2 could generate a false sensation of security among dentists, care staff, and patients, which could subsequently lead to a groundless reduction in the use of known effective measures for the prevention of infection and dissemination of COVID-19 during aerosol-generating procedures resulting in an increment in contagion rates.

## Acknowledgments

The authors are grateful to Donna Pringle for reviewing the language and style.

## Author contributions

AHV and DA conceived the study. All authors contributed to the design of the study. DA and AHV were involved in planning, supervised, and validated the work. DC executed the searches and de-duplicated the references. For the review, AHV and ABP contributed to the data screening, data extraction, and assessment of study quality. AHV, ABP, and DA contributed to the analysis of the results and to the writing of the manuscript. All authors contributed to the preparation of tables and figures. All authors read and approved the final manuscript.

## Sources of Finance

None.

