## Appendix Table 1 for "Mouth-rinses and SARS-CoV-2 viral load in saliva: A living systematic review"

### APPENDIX FILES

**Appendix References 1.** List of included studies in this living systematic review.

Gottsauner MJ, Michaelides I, Schmidt B, Scholz KJ, Buchalla W, Widbiller M, Hitzenbichler F, Ettl T, Reichert TE, Bohr C, et al. 2020. A prospective clinical pilot study on the effects of a hydrogen peroxide mouthrinse on the intraoral viral load of SARS-CoV-2. Clin Oral Investig. 24(10):3707–3713. doi:10.1007/s00784-020-03549-1.

Yoon JG, Yoon J, Song JY, Yoon SY, Lim CS, Seong H, Noh JY, Cheong HJ, Kim WJ. 2020. Clinical Significance of a High SARS-CoV-2 Viral Load in the Saliva. J Korean Med Sci. 35(20):e195. doi:10.3346/jkms.2020.35.e195.

Martínez Lamas L, Diz Dios P, Pérez Rodríguez MT, Del Campo Pérez V, Cabrera Alvargonzalez JJ, López Domínguez AM, Fernandez Feijoo J, Diniz Freitas M, Limeres Posse J. 2020 Jul 29. Is povidone iodine mouthwash effective against SARS-CoV-2? First in vivo tests. Oral Dis.:odi.13526. doi:10.1111/odi.13526.

Seneviratne CJ, Balan P, Ko KKK, Udawatte NS, Lai D, Ng DHL, Venkatachalam I, Lim KS, Ling ML, Oon L, et al. 2020 Dec 14. Efficacy of commercial mouth-rinses on SARS-CoV-2 viral load in saliva: randomized control trial in Singapore. Infection. doi:10.1007/s15010-020-01563-9.

**Appendix Table 1.** Search Strategies.

| **Database** | **PubMed**<br><br>**Date:** February 26, 2021 | | **Results** |
|---|---|---|---|
| **Search Strategy** | #1 | SARS-CoV-2[Mesh] | 47,373 |
| | #2 | COVID-19[Mesh] | 60,070 |
| | #3 | Corona Virus[tiab] | 1,705 |
| | #4 | COVID-19[tiab] | 93,344 |
| | #5 | COVID19*[tiab] | 89,688 |
| | #6 | 2019-nCoV[tiab] | 1,485 |
| | #7 | SARS-CoV-2[tiab] | 33,769 |
| | #8 | SARS-CoV2[tiab] | 1,508 |
| | #9 | (Pneumonia[tiab] AND Wuhan[tiab] AND 2019[tiab]) | 1,025 |

| | | | |
|---|---|---|---|
| | #10 | Coronavir*[tiab] | 54,420 |
| | #11 | Coronovir*[tiab] | 50 |
| | #12 | Virus Corona[tiab] | 2,247 |
| | #13 | Corono Virus[tiab] | 1 |
| | #14 | HCov*[tiab] | 937 |
| | #15 | CV19*[tiab] | 17 |
| | #16 | CV-19[tiab] | 2 |
| | #17 | N-Cov[tiab] | 8 |
| | #18 | #1 OR #2 OR #3 OR #4 OR #5 OR #6 OR #7 OR #8 OR #9 OR #10 OR #11 OR #12 OR #13 OR #14 OR #15 OR #16 OR #17 | 117,784 |
| | #19 | Mouthwashes[Mesh] | 7,001 |
| | #20 | Mouth Rinse*[tiab] | 1,030 |
| | #21 | Mouthrinse*[tiab] | 1,693 |
| | #22 | Mouth Bath*[tiab] | 7 |
| | #23 | Mouthbath*[tiab] | 1 |
| | #24 | Mouth Wash*[tiab] | 321 |
| | #25 | Mouthwash*[tiab] | 2,859 |
| | #26 | Oral Rinse*[tiab] | 809 |
| | #27 | Oralrinse*[tiab] | 15 |
| | #28 | Oral Bath*[tiab] | 1,659 |
| | #29 | Oral Wash*[tiab] | 117 |
| | #30 | Oralwash*[tiab] | 1 |
| | #31 | Dental Rinse*[tiab] | 21 |
| | #32 | Dentalrinse*[tiab] - Schema: all | 0 |
| | #33 | Dentalrinse*[tiab] | 0 |
| | #34 | Dental Bath*[tiab] | 896 |
| | #35 | Dental Wash*[tiab] | 2,620 |
| | #36 | #19 OR #20 OR #21 OR #22 OR #23 OR #24 OR #25 OR #26 OR #27 OR #28 OR #29 OR #30 OR #31 OR #32 OR #33 OR #34 OR #35 | 14,351 |
| | #37 | #18 AND #36 | 115 |

| Database | CINAHL<br><br>**Date:** February 26, 2021 | | **Results** |
|---|---|---|---|
| **Search Strategy** | S1 | (MH "COVID-19") | 13,413 |
| | S2 | (MH "Severe Acute Respiratory Syndrome") | 2,459 |
| | S3 | TI (Coron* N1 Virus) OR AB (Coron* N1 Virus) | 395 |
| | S4 | TI COVID-19 OR AB COVID-19 | 31,631 |
| | S5 | TI COVID19* OR AB COVID19* | 189 |
| | S6 | TI 2019-nCoV OR AB 2019-nCoV | 229 |
| | S7 | TI SARS-CoV-2 OR AB SARS-CoV-2 | 5,013 |
| | S8 | TI SARS-CoV2 OR AB SARS-CoV2 | 163 |
| | S9 | TI SARSCoV2 OR AB SARSCoV2 | 6 |
| | S10 | TI (Pneumonia AND Wuhan AND 2019) OR AB (Pneumonia AND Wuhan AND 2019) | 200 |
| | S11 | TI Coronavir* OR AB Coronavir* | 12,582 |
| | S12 | TI HCov* OR AB HCov* | 77 |
| | S13 | TI CV19* OR AB CV19* | 1 |
| | S14 | TI CV-19* OR AB CV-19* | 1 |
| | S15 | TI N-Cov OR AB N-Cov | 1 |
| | S16 | S1 OR S2 OR S3 OR S4 OR S5 OR S6 OR S7 OR S8 OR S9 OR S10 OR S11 OR S12 OR S13 OR S14 OR S15 | 25,915 |
| | S17 | (MH "Mouthwashes+") | 2,478 |
| | S18 | TI (Mouth N1 Rinse*) OR AB (Mouth N1 Rinse*) | 481 |
| | S19 | TI Mouthrinse* OR AB Mouthrinse* | 460 |
| | S20 | TI (Mouth N1 Bath*) OR AB (Mouth N1 Bath*) | 7 |
| | S21 | TI Mouthbath* OR AB Mouthbath* | 0 |
| | S22 | TI (Mouth N1 Wash*) OR AB (Mouth N1 Wash*) | 71 |
| | S23 | TI Mouthrinse* OR AB Mouthrinse* | 357 |
| | S24 | TI (Mouth N1 Bath*) OR AB (Mouth N1 Bath*) | 7 |
| | S25 | TI Mouthbath* OR AB Mouthbath* | 0 |
| | S26 | TI (Mouth N1 Wash*) OR AB (Mouth N1 Wash*) | 66 |
| | S27 | TI Mouthwash* OR AB Mouthwash* | 712 |

| | | | |
|---|---|---|---|
| | S28 | TI (Oral N1 Rinse*) OR AB (Oral N1 Rinse*) | 283 |
| | S29 | TI Oralrinse* OR AB Oralrinse* | 0 |
| | S30 | TI (Oral N1 Bath*) OR AB (Oral N1 Bath*) | 10 |
| | S31 | TI (Oral N1 Wash*) OR AB (Oral N1 Wash*) | 34 |
| | S32 | TI Oralwash OR AB Oralwash* | 0 |
| | S33 | TI (Dental N1 Rinse*) OR AB (Dental N1 Rinse*) | 10 |
| | S34 | TI Dentalrinse* OR AB Dentalrinse* | 0 |
| | S35 | TI (Dental N1 Bath*) OR AB (Dental N1 Bath*) | 2 |
| | S36 | TI (Dental N1 Wash*) OR AB (Dental N1 Wash*) | 20 |
| | S37 | S17 OR S18 OR S19 OR S20 OR S21 OR S22 OR S23 OR S24 OR S25 OR S26 OR S27 OR S28 OR S29 OR S30 OR S31 OR S32 OR S33 OR S34 OR S35 OR S36 | 2,301 |
| | S38 | S16 AND S37 | 24 |

| **Database** | **Cochrane Library**<br><br>**Date:** February 26, 2021 | | **Results** |
|---|---|---|---|
| **Search Strategy** | #1 | MeSH descriptor: [SARS-CoV-2] explode all trees | 201 |
| | #2 | MeSH descriptor: [COVID-19] explode all trees | 251 |
| | #3 | (Coron* NEAR/1 Virus):ti,ab,kw | 187 |
| | #4 | COVID-19:ti,ab,kw | 4186 |
| | #5 | COVID19:ti,ab,kw | 257 |
| | #6 | "2019-nCoV":ti,ab,kw | 10 |
| | #7 | "SARS-CoV-2":ti,ab,kw | 186 |
| | #8 | "SARS-CoV2":ti,ab,kw | 51 |
| | #9 | SARSCoV2:ti,ab,kw | 162 |
| | #10 | (Pneumonia AND Wuhan AND 2019):ti,ab,kw | 86 |
| | #11 | Coronavir*:ti,ab,kw | 2527 |
| | #12 | Coronovir*:ti,ab,kw | 1 |
| | #13 | HCov*:ti,ab,kw | 15 |
| | #14 | CV19*:ti,ab,kw | 0 |
| | #15 | CV-19*:ti,ab,kw | 12 |

| | | | |
|---|---|---|---|
| | #16 | "N-Cov":ti,ab,kw | 29 |
| | #17 | #1 OR #2 OR #3 OR #4 OR #5 OR #6 OR #7 OR #8 OR #9 OR #10 OR #11 OR #12 OR #13 OR #14 OR #15 OR #16 | 4490 |
| | #18 | MeSH descriptor: [Mouthwashes] explode all trees | 1659 |
| | #19 | (Mouth NEAR/1 Rinse*):ti,ab,kw | 766 |
| | #20 | Mouthrinse*:ti,ab,kw | 1196 |
| | #21 | (Mouth NEAR/1 Bath*):ti,ab,kw | 2 |
| | #22 | Mouthbath*:ti,ab,kw | 0 |
| | #23 | (Mouth NEAR/1 Wash*):ti,ab,kw | 275 |
| | #24 | Mouthwash*:ti,ab,kw | 3041 |
| | #25 | (Oral NEAR/1 Rinse*):ti,ab,kw | 363 |
| | #26 | Oralrinse*:ti,ab,kw | 3 |
| | #27 | (Oral NEAR/1 Bath*):ti,ab,kw | 2 |
| | #28 | (Oral NEAR/1 Wash*):ti,ab,kw | 37 |
| | #29 | Oralwash*:ti,ab,kw | 0 |
| | #30 | (Dental NEAR/1 Rinse*):ti,ab,kw | 32 |
| | #31 | Dentalrinse*:ti,ab,kw | 0 |
| | #32 | (Dental NEAR/1 Bath*):ti,ab,kw | 0 |
| | #33 | (Dental NEAR/1 Wash*):ti,ab,kw | 3 |
| | #34 | #18 OR #19 OR #20 OR #21 OR #22 OR #23 OR #24 OR #25 OR #26 OR #27 OR #28 OR #29 OR #30 OR #31 OR #32 OR #33 | 4391 |
| | #35 | #17 AND #34 | 32 |

| **Database** | **Embase**<br><br>**Date:** February 26, 2021 | | **Results** |
|---|---|---|---|
| **Search Strategy** | #1 | 'severe acute respiratory syndrome coronavirus 2'/exp | 22,785 |
| | #2 | 'coronavirus disease 2019'/exp | 87,854 |
| | #3 | (coron* NEAR/1 virus):ti,ab | 1,850 |
| | #4 | 'covid 19':ti,ab | 85,652 |
| | #5 | covid19*:ti,ab | 1,161 |
| | #6 | '2019 ncov':ti,ab | 1,146 |

| | | | |
|---|---|---|---|
| | #7 | 'sars-cov-2':ti,ab | 26,761 |
| | #8 | 'sars-cov2':ti,ab | 1,347 |
| | #9 | 'sarscov2':ti,ab | 48 |
| | #10 | pneumonia:ti,ab AND wuhan:ti,ab AND 2019:ti,ab | 1,010 |
| | #11 | coronavir*:ti,ab | 48,260 |
| | #12 | coronovir*:ti,ab | 51 |
| | #13 | hcov*:ti,ab | 963 |
| | #14 | cv19*:ti,ab | 18 |
| | #15 | 'cv 19*':ti,ab | 122 |
| | #16 | 'n cov':ti,ab | 24 |
| | #17 | #1 OR #2 OR #3 OR #4 OR #5 OR #6 OR #7 OR #8 OR #9 OR #10 OR #11 OR #12 OR #13 OR #14 OR #15 OR #16 | 119,174 |
| | #18 | 'mouthwash'/exp | 4,889 |
| | #19 | (mouth NEAR/1 rinse*):ti,ab | 1,176 |
| | #20 | mouthrinse*:ti,ab | 1,583 |
| | #21 | (mouth NEAR/1 bath*):ti,ab | 12 |
| | #22 | mouthbath*:ti,ab | 0 |
| | #23 | (mouth NEAR/1 wash*):ti,ab | 490 |
| | #24 | mouthwash*:ti,ab | 3,447 |
| | #25 | (oral NEAR/1 rinse*):ti,ab | 964 |
| | #26 | oralrinse*:ti,ab | 0 |
| | #27 | (oral NEAR/1 bath*):ti,ab | 21 |
| | #28 | (oral NEAR/1 wash*):ti,ab | 208 |
| | #29 | oralwash*:ti,ab | 1 |
| | #30 | (dental NEAR/1 rinse*):ti,ab | 32 |
| | #31 | dentalrinse*:ti,ab | 0 |
| | #32 | (dental NEAR/1 bath*):ti,ab | 0 |
| | #33 | (dental NEAR/1 wash*):ti,ab | 24 |
| | #34 | #18 OR #19 OR #20 OR #21 OR #22 OR #23 OR #24 OR #25 OR #26 OR #27 OR #28 OR #29 OR #30 OR #31 OR #32 OR #33 | 9,061 |
| | #35 | #17 AND #34 | 109 |

| **Database** | **Scopus**<br><br>**Date:** February 26, 2021 | | **Results** |
|---|---|---|---|
| **Search Strategy** | #1 | TITLE-ABS-KEY ("SARS-CoV-2") | 36,361 |
| | #2 | TITLE-ABS-KEY ("COVID-19") | 103,116 |
| | #3 | TITLE-ABS-KEY ("Corona Virus") | 2,459 |
| | #4 | TITLE-ABS-KEY (COVID19*) | 1,547 |
| | #5 | TITLE-ABS-KEY ("2019-nCoV") | 1,610 |
| | #6 | TITLE-ABS-KEY ("SARS-CoV2") | 1,414 |
| | #7 | TITLE-ABS-KEY (Pneumonia AND Wuhan AND 2019) | 2,555 |
| | #8 | TITLE-ABS-KEY (Coronavir*) | 102,097 |
| | #9 | TITLE-ABS-KEY (Coronovir*) | 101 |
| | #10 | TITLE-ABS-KEY ("Virus Corona") | 19 |
| | #11 | TITLE-ABS-KEY ("Corono Virus") | 3 |
| | #12 | TITLE-ABS-KEY (HCov*) | 910 |
| | #13 | TITLE-ABS-KEY (CV19*) | 20 |
| | #14 | TITLE-ABS-KEY ("CV-19") | 52 |
| | #15 | TITLE-ABS-KEY ("N-Cov") | 36 |
| | #16 | #1 OR #2 OR #3 OR #4 OR #5 OR #6 OR #7 OR #8 OR #9 OR #10 OR #11 OR #12 OR #13 OR #14 OR #15 | 138,151 |
| | #17 | TITLE-ABS-KEY ("Mouthwashes") | 8,756 |
| | #18 | TITLE-ABS-KEY (Mouth Rinse*) | 3,123 |
| | #19 | TITLE-ABS-KEY (Mouthrinse*) | 1,809 |
| | #20 | TITLE-ABS-KEY (Mouth Bath*) | 1,386 |
| | #21 | TITLE-ABS-KEY (Mouthbath*) | 0 |
| | #22 | TITLE-ABS-KEY (Mouth Wash*) | 3,080 |
| | #23 | TITLE-ABS-KEY (Mouthwash*) | 8,776 |
| | #24 | TITLE-ABS-KEY (Oral Rinse*) | 3,106 |
| | #25 | TITLE-ABS-KEY (Oralrinse*) | 0 |
| | #26 | TITLE-ABS-KEY (Oral Bath*) | 1,627 |

| | | | |
|---|---|---|---|
| | #27 | TITLE-ABS-KEY (Oral Wash*) | 9,826 |
| | #28 | TITLE-ABS-KEY (Oralwash*) | 1 |
| | #29 | TITLE-ABS-KEY (Dental Rinse*) | 4,500 |
| | #30 | TITLE-ABS-KEY (Dentalrinse*) | 0 |
| | #31 | TITLE-ABS-KEY (Dental Bath*) | 843 |
| | #32 | TITLE-ABS-KEY (Dental Wash*) | 2,894 |
| | #33 | #17 OR #18 OR #19 OR #20 OR #21 OR #22 OR #23 OR #24 OR #25 OR #26 OR #27 OR #28 OR #29 OR #30 OR #31 OR #32 | 29,926 |
| | #34 | #16 AND #33 | 259 |

| **Database** | **Dentistry & Oral Sciences Source**<br><br>**Date:** February 26, 2021 | | **Results** |
|---|---|---|---|
| **Search Strategy** | S1 | (MH "COVID-19") | 13,413 |
| | S2 | (MH "Severe Acute Respiratory Syndrome") | 2,459 |
| | S3 | TI (Coron* N1 Virus) OR AB (Coron* N1 Virus) | 395 |
| | S4 | TI COVID-19 OR AB COVID-19 | 31,631 |
| | S5 | TI COVID19* OR AB COVID19* | 189 |
| | S6 | TI 2019-nCoV OR AB 2019-nCoV | 229 |
| | S7 | TI SARS-CoV-2 OR AB SARS-CoV-2 | 5,013 |
| | S8 | TI SARS-CoV2 OR AB SARS-CoV2 | 163 |
| | S9 | TI SARSCoV2 OR AB SARSCoV2 | 6 |
| | S10 | TI (Pneumonia AND Wuhan AND 2019) OR AB (Pneumonia AND Wuhan AND 2019) | 200 |
| | S11 | TI Coronavir* OR AB Coronavir* | 12,582 |
| | S12 | TI HCov* OR AB HCov* | 77 |
| | S13 | TI CV19* OR AB CV19* | 1 |
| | S14 | TI CV-19* OR AB CV-19* | 1 |
| | S15 | TI N-Cov OR AB N-Cov | 1 |
| | S16 | S1 OR S2 OR S3 OR S4 OR S5 OR S6 OR S7 OR S8 OR S9 OR S10 OR S11 OR S12 OR S13 OR S14 OR S15 | 25,915 |
| | S17 | (MH "Mouthwashes+") | 2,478 |

| | | | |
|---|---|---|---|
| | S18 | TI (Mouth N1 Rinse*) OR AB (Mouth N1 Rinse*) | 481 |
| | S19 | TI Mouthrinse* OR AB Mouthrinse* | 460 |
| | S20 | TI (Mouth N1 Bath*) OR AB (Mouth N1 Bath*) | 7 |
| | S21 | TI Mouthbath* OR AB Mouthbath* | 0 |
| | S22 | TI (Mouth N1 Wash*) OR AB (Mouth N1 Wash*) | 71 |
| | S23 | TI Mouthrinse* OR AB Mouthrinse* | 357 |
| | S24 | TI (Mouth N1 Bath*) OR AB (Mouth N1 Bath*) | 7 |
| | S25 | TI Mouthbath* OR AB Mouthbath* | 0 |
| | S26 | TI (Mouth N1 Wash*) OR AB (Mouth N1 Wash*) | 66 |
| | S27 | TI Mouthwash* OR AB Mouthwash* | 712 |
| | S28 | TI (Oral N1 Rinse*) OR AB (Oral N1 Rinse*) | 283 |
| | S29 | TI Oralrinse* OR AB Oralrinse* | 0 |
| | S30 | TI (Oral N1 Bath*) OR AB (Oral N1 Bath*) | 10 |
| | S31 | TI (Oral N1 Wash*) OR AB (Oral N1 Wash*) | 34 |
| | S32 | TI Oralwash OR AB Oralwash* | 0 |
| | S33 | TI (Dental N1 Rinse*) OR AB (Dental N1 Rinse*) | 10 |
| | S34 | TI Dentalrinse* OR AB Dentalrinse* | 0 |
| | S35 | TI (Dental N1 Bath*) OR AB (Dental N1 Bath*) | 2 |
| | S36 | TI (Dental N1 Wash*) OR AB (Dental N1 Wash*) | 20 |
| | S37 | S17 OR S18 OR S19 OR S20 OR S21 OR S22 OR S23 OR S24 OR S25 OR S26 OR S27 OR S28 OR S29 OR S30 OR S31 OR S32 OR S33 OR S34 OR S35 OR S36 | 2,301 |
| | S38 | S16 AND S37 | 24 |

| **Database** | **LILACS**<br><br>**Date:** February 26, 2021 | | **Results** |
|---|---|---|---|
| **Search Strategy** | #1 | (MH SARS-CoV-2 OR MH COVID-19 OR Corona OR COVID-19 OR COVID19$ OR 2019-nCoV OR SARS-CoV-2 OR SARS-CoV2 OR Coronavir$ OR Coronovir$ OR HCov$ OR CV19$ OR CV-19 OR N-Cov) AND (MH Mouthwashes OR Mouthwas$ OR Mouthrinse$ OR Mouthbath$ OR Rinse$ OR Enjuague$ OR Enxague) [Words] | 18 |

| **Database** | **medRxiv AND bioRxiv** | | **Results** |
|---|---|---|---|

| | | | |
|---|---|---|---|
| | **Date:** February 26, 2021 | | |
| **Search Strategy** | #1 | "Mouthwashes AND Covid" | 24 |

**Appendix Checklist 1.** The PRISMA Statement[1].

| Section/topic | # | Checklist item | Reported on page # |
|---|---|---|---|
| **TITLE** | | | |
| Title | 1 | Identify the report as a systematic review, meta-analysis, or both. | 1 |
| **ABSTRACT** | | | |
| Structured summary | 2 | Provide a structured summary including, as applicable: background; objectives; data sources; study eligibility criteria, participants, and interventions; study appraisal and synthesis methods; results; limitations; conclusions and implications of key findings; systematic review registration number. | 2 |
| **INTRODUCTION** | | | |
| Rationale | 3 | Describe the rationale for the review in the context of what is already known. | 3 |
| Objectives | 4 | Provide an explicit statement of questions being addressed with reference to participants, interventions, comparisons, outcomes, and study design (PICOS). | 3 |
| **METHODS** | | | |
| Protocol and registration | 5 | Indicate if a review protocol exists, if and where it can be accessed (e.g., Web address), and, if available, provide registration information including registration number. | 4 |
| Eligibility criteria | 6 | Specify study characteristics (e.g., PICOS, length of follow-up) and report characteristics (e.g., years considered, language, publication status) used as criteria for eligibility, giving rationale. | 4 |
| Information sources | 7 | Describe all information sources (e.g., databases with dates of coverage, contact with study authors to identify additional studies) in the search and date last searched. | 4 |
| Search | 8 | Present full electronic search strategy for at least one database, including any limits used, such that it could be repeated. | 4 |
| Study selection | 9 | State the process for selecting studies (i.e., screening, eligibility, included in systematic review, and, if applicable, included in the meta-analysis). | 4 |
| Data collection process | 10 | Describe method of data extraction from reports (e.g., piloted forms, independently, in duplicate) and any processes for obtaining and confirming data from investigators. | 4-5 |
| Data items | 11 | List and define all variables for which data were sought (e.g., PICOS, funding sources) and any assumptions and simplifications made. | 5 |
| Risk of bias in individual studies | 12 | Describe methods used for assessing risk of bias of individual studies (including specification of whether this was done at the study or outcome level), and how this information is to be used in any data synthesis. | 5 |
| Summary measures | 13 | State the principal summary measures (e.g., risk ratio, difference in means). | 5 |

| | | | |
|---|---|---|---|
| Synthesis of results | 14 | Describe the methods of handling data and combining results of studies, if done, including measures of consistency (e.g., $I^2$) for each meta-analysis. | NA |



| Section/topic | # | Checklist item | Reported on page # |
|---|---|---|---|
| Risk of bias across studies | 15 | Specify any assessment of risk of bias that may affect the cumulative evidence (e.g., publication bias, selective reporting within studies). | 5 |
| Additional analyses | 16 | Describe methods of additional analyses (e.g., sensitivity or subgroup analyses, meta-regression), if done, indicating which were pre-specified. | NA |
| **RESULTS** | | | |
| Study selection | 17 | Give numbers of studies screened, assessed for eligibility, and included in the review, with reasons for exclusions at each stage, ideally with a flow diagram. | 6 |
| Study characteristics | 18 | For each study, present characteristics for which data were extracted (e.g., study size, PICOS, follow-up period) and provide the citations. | 6 |
| Risk of bias within studies | 19 | Present data on risk of bias of each study and, if available, any outcome level assessment (see item 12). | 9 |
| Results of individual studies | 20 | For all outcomes considered (benefits or harms), present, for each study: (a) simple summary data for each intervention group (b) effect estimates and confidence intervals, ideally with a forest plot. | 6-9 |
| Synthesis of results | 21 | Present results of each meta-analysis done, including confidence intervals and measures of consistency. | 6-9 |
| Risk of bias across studies | 22 | Present results of any assessment of risk of bias across studies (see Item 15). | 9 |
| Additional analysis | 23 | Give results of additional analyses, if done (e.g., sensitivity or subgroup analyses, meta-regression [see Item 16]). | NA |
| **DISCUSSION** | | | |
| Summary of evidence | 24 | Summarize the main findings including the strength of evidence for each main outcome; consider their relevance to key groups (e.g., healthcare providers, users, and policy makers). | 10 |
| Limitations | 25 | Discuss limitations at study and outcome level (e.g., risk of bias), and at review-level (e.g., incomplete retrieval of identified research, reporting bias). | 12 |
| Conclusions | 26 | Provide a general interpretation of the results in the context of other evidence, and implications for future research. | 12-13 |
| **FUNDING** | | | |
| Funding | 27 | Describe sources of funding for the systematic review and other support (e.g., supply of data); role of funders for the systematic review. | 1 |

[1] Moher D, Liberati A, Tetzlaff J, Altman DG, The PRISMA Group (2009). Preferred Reporting Items for Systematic Reviews and Meta-Analyses: The PRISMA Statement. PLoS Med 6(6): e1000097. doi:10.1371/journal.pmed1000097
